# Both HFrEF and HFpEF Should be Included in Calculating CHA2DS2–VASc score: a Taiwanese Longitudinal Cohort

**DOI:** 10.1101/2023.10.25.23297579

**Authors:** Chien-Chien Cheng, Pang-Shuo Huang, Jien-Jiun Chen, Fu-Chun Chiu, Sheng-Nan Chang, Yi-Chih Wang, Cho-Kai Wu, Juey-Jen Hwang, Chia-Ti Tsai

**Affiliations:** National Taiwan University College of Medicine, Taipei, Taiwan; Division of Cardiology, Department of Internal Medicine, National Taiwan University Hospital, Taipei, Taiwan; Division of Cardiology, Department of Internal Medicine, National Taiwan University Hospital Yun-Lin Branch, Yun-Lin, Taiwan; Graduate Institute of Clinical Medicine, College of Medicine, National Taiwan University, Taipei, Taiwan; Cardiovascular Center, National Taiwan University Hospital, Taipei, Taiwan

**Keywords:** Atrial fibrillation, HFrEF, HFpEF, CHA2DS2–VASc score, Ischemic stroke

## Abstract

**Background:** Congestive heart failure (CHF) in the context of AF-related stroke risk, was coined when it mainly referred to patients with left ventricular systolic dysfunction (HFrEF). However, the term now encompasses patients with preserved ejection fraction (HFpEF) as well. Given this change, it becomes essential to investigate the variation in stroke risk between atrial fibrillation (AF) patients with HFpEF and HFrEF for enhancing risk assessment and subsequent management strategies.

**Methods:** In a longitudinal study utilizing the National Taiwan University Hospital Integrated Medical Database (iMED), 8358 patients with AF were followed up for 10 years from January 2010 to December 2020 (mean follow-up 3.76 years). The study evaluated the risk of ischemic stroke, using Cox models adjusted for potential risk factors of AF-related stroke.

**Results:** Comparing AF patients of different CHF subgroups, HFpEF patients had a higher mean CHA2DS2–VASc score (4.08±1.502 vs. 3.83±1.571, p<0.001) and a higher risk of stroke during follow-up (HR 1.151 (1.013-1.308), p=0.031). In contrast, patients with HFrEF had a higher prevalence of myocardial infarction (MI) and coronary artery disease (CAD). After adjusting for other risk factors, there was no significant difference in the risk of new-onset stroke between HFpEF and HFrEF patients (HR 1.001 (0.877-1.142), p=0.994).

**Conclusion:** After adjusting for other risk factors of stroke, both HFpEF and HFrEF were found to have a similar risk of stroke in AF patients. Therefore, it is important to extend the criteria for “C” in the CHA2DS2–VASc score to include HFpEF patients. Prior to multivariable adjustment, HFpEF patients had a higher risk than those with HFrEF, likely due to their higher CHA2DS2–VASc score, indicating a greater prevalence of stroke-related comorbidities.

## Introduction

Atrial fibrillation (AF) has become the most prevalent cardiac arrhythmic disease, and its prevalence is continuing to rise, in part due to the aging population. AF is associated with multiple comorbidities, including up to five-fold increased risk of ischemic stroke(1). Congestive heart failure (CHF) has been a growing concern due to an aging population, with an annual incidence of 177 per 100,000 people in Taiwan (2). Potential risk factors of CHF include coronary artery disease (CAD), valvular heart disease, myocardial infarction (MI), and AF. Frequently coexisting, CHF has also been known to predispose AF, and vice versa, further acting as a risk factor of stroke development in this population.

When predicting stroke incidence in AF patients, current guidelines recommend utilizing the CHA2DS2–VASc score, a convenient risk stratification instrument for stroke risk in AF patients (3, 4). It is widely utilized as it incorporates mostly known patient history and characteristics, including CHF. Nevertheless, as the understanding of CHF advances, the term is no longer limited to patients with reduced ejection fraction (HFrEF), and is now defined as a clinical syndrome with classic signs or symptoms caused by a cardiac abnormality(5). Although this definition includes a subset of patients with preserved ejection fraction (HFpEF), an entity commonly defined as LVEF≥50%, HFpEF itself is not sufficient to fulfill the “C” criterion in current AF management guidelines. Interestingly, according to a survey conducted by the European Heart Rhythm Association (EHRA) Scientific Initiatives Committee on the interpretation of CHA2DS2-VASc score components in clinical practice, the “C” component was among those difficult to score. This reflects a knowledge gap in current scoring systems, as inconsistencies exist for similar terminology. Therefore, further exploration of the impact of HFpEF is needed for clarity of diagnosis definition.

Atrial fibrillation has been shown to be more prevalent in HFpEF patients (6, 7), and the two phenotypes to some extent have an interdependent relationship clinically and epidemiologically (8). Nevertheless, studies have also shown no difference in the development of HFrEF versus HFpEF in AF patients (9), and the impact of ejection fraction and occurrence of ischemic stroke is controversial.

The CHA2DS2–VASc score was developed prior to the introduction of a broadened scope of heart failure subtypes and the role of HFpEF in this setting is ambiguous(10). Patients with HFpEF have been shown to have a higher score, and when compared to HFrEF patients, are at a greater risk of developing stroke and systemic embolism (11). Despite the aforementioned advances, the impact of coexisting risk factors has not been clarified. This study aims to delineate the impact of HFpEF versus HFrEF with patient comorbidities taken into consideration.

## Methods

### Study population

This retrospective, cross-sectional study took place at National Taiwan University Hospital (NTUH), a tertiary medical center in Taiwan. Data from both the NTUH Integrated Medical Database (NTUH-IMD) and reimbursement claim records from the Taiwan national health insurance program at NTUH were used for the study. To ensure data accuracy and integrity, chart reviews were conducted as necessary to supplement the information obtained from the NTUH-IMD. The study period spanned from January 2010 to December 2020.

The study protocol received approval from the Research Ethics Committee of the NTUH, and informed consent was waived given the retrospective nature of the study. Patient data with any identifying information was accessible only to the investigators and kept confidential in accordance with the approved protocol 200911002R and 202011019RINC. During the study period, an AF cohort was formed using a common criterion, which required at least three outpatient claims within a year or at least one inpatient claim coded for AF (ICD9-CM codes 427.31) in the first three or five diagnostic codes, respectively. No restrictions were imposed on age or other parameters in this cohort. HFpEF was defined as patients with a left ventricular ejection fraction (LVEF) of 55% or higher, while HFrEF was defined as patients with an LVEF below 55%.

8358 patients with atrial fibrillation (AF) were followed up for 10 years (mean follow-up 3.76 years, median follow-up time 3.21 years). Among these patients, 4654 (55.7%) had no history of CHF, 2374 (28.4%) had HFpEF, and 1330 (15.9%) had HFrEF.

The study aimed to assess the risk of thromboembolic events, specifically ischemic stroke, in the HFpEF population and compare it with traditional HFrEF populations among AF patients.

### Statistical analysis

Continuous data were presented as mean ± standard deviation (SD). Independent two-sample Student’s t-test was used for comparing continuous variables, while the chi-square test was employed for categorical variables. The time to the first event of ischemic stroke was illustrated using the Kaplan–Meier survival function estimation. Survival curve differences were tested using log-rank statistics.

For identifying predictors (specifically HFpEF and HFrEF and other components of CHA2DS2–VASc score) of incident ischemic stroke, multivariable Cox proportional-hazards regression analysis was performed, and hazard ratios (HRs) with corresponding 95% confidence intervals (CIs) were calculated.

All statistical tests were two-sided, and a significance level of p < 0.05 was considered statistically significant. The statistical analyses were carried out using SPSS 25.0 (IBM Corp, Armonk, NY).

## Results

### Baseline characteristics

The baseline characteristics of the study participants are shown in Table 1. Comparing AF patients without CHF, with HFpEF, or with HFrEF, we found that patients with HFpEF were oldest, more female, and had a higher prevalence of previous stroke and hypertension. They also had a higher mean CHA2DS2–VASc score (4.08±1.502 vs. 3.83±1.571 vs. 2.52±1.513, p<0.001). In contrast, patients with HFrEF were younger and had a higher prevalence of MI and CAD, likely due to an ischemic cardiomyopathy etiology.

**Table 1.**
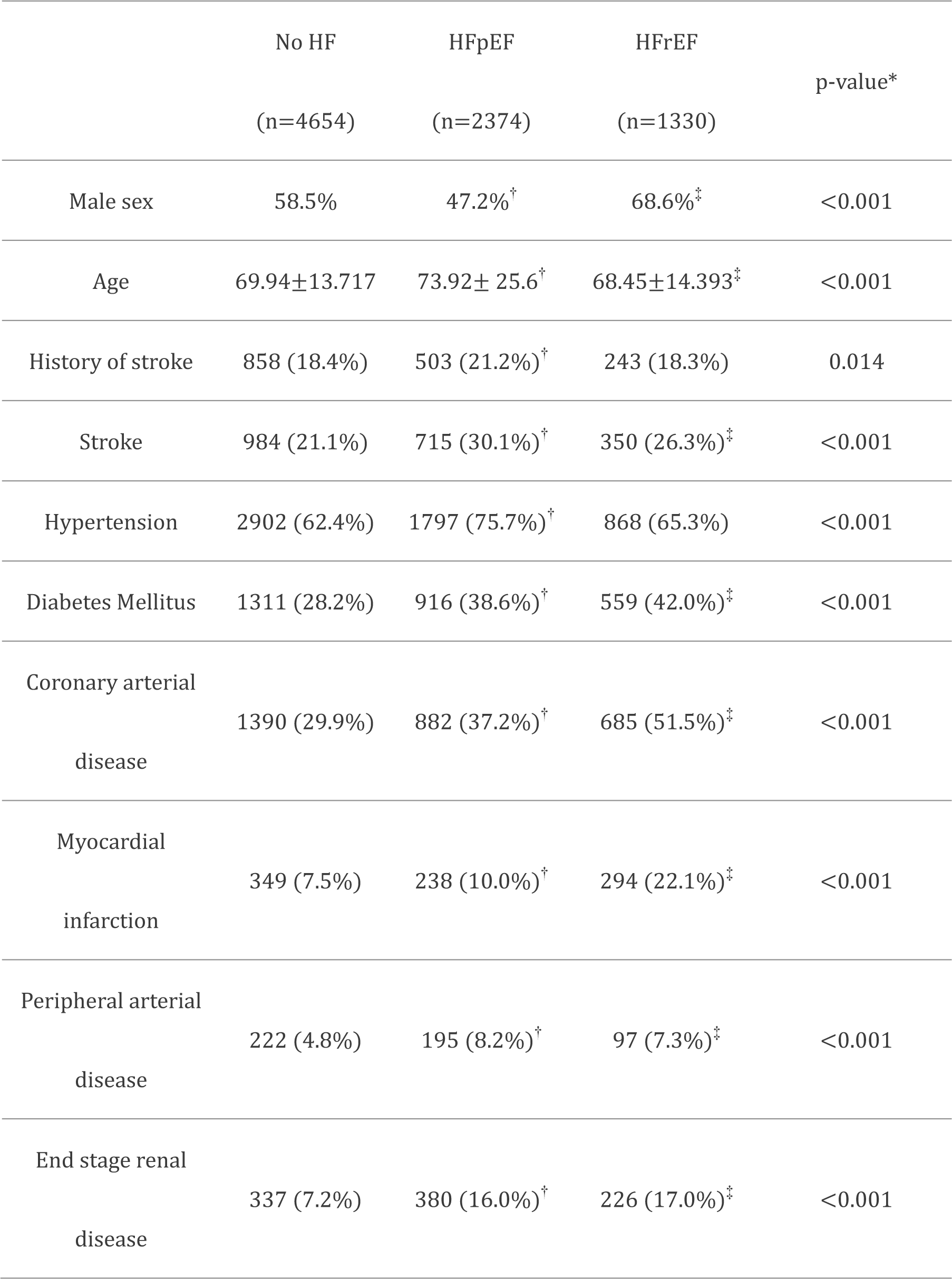
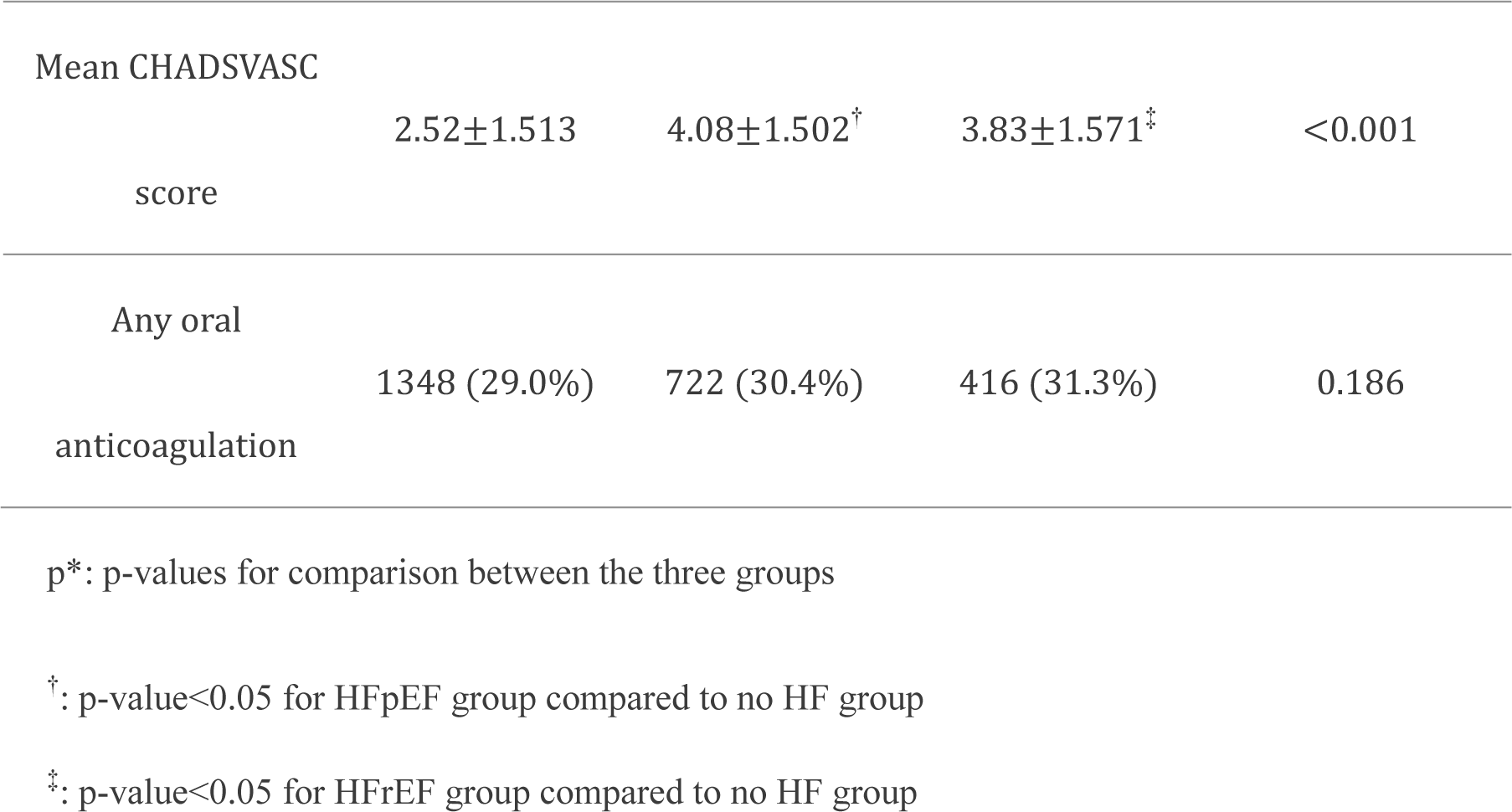
Comparisons of clinical characteristics of patients in the different HF groups according to ejection fraction.

### HFpEF patients had a higher risk of developing ischemic stroke when compared to HFrEF patients during long-term follow-up

A total of 2049 new-onset strokes occurred during long-term follow-up (Table 1). Comparing patients with CHF to those without (Table 2), as expected, patients with CHF were more likely to have an attack of ischemic stroke (HR 1.358 (1.245-1.481), p<0.001). Further stratifying these patients according to ejection fraction, we found that patients with HFpEF had a higher risk of stroke than those with HFrEF (HR 1.425 vs 1.238). The risk of stroke development was significantly higher in HFpEF patients (HR 1.151 (1.013-1.308), p=0.031, with HFrEF as reference).

**Table 2.** Hazard ratios (HRs) for heart failure as a whole (CHF) vs no CHF, heart failure with preserved ejection fraction (HFpEF) vs no HF and heart failure with reduced ejection fraction (HFrEF) vs. no HF for stroke event during long term follow up.

|  | HR | 95.0% CI for HR |  | p-value |
| --- | --- | --- | --- | --- |
|  |  | Lower | Upper |  |
| CHF vs no CHF | 1.358 | 1.245 | 1.481 | <0.001 |
| HFpEF vs no CHF | 1.425 | 1.294 | 1.569 | <0.001 |
| HFrEF vs no CHF | 1.238 | 1.096 | 1.399 | 0.001 |
| HFpEF vs HFrEF | 1.151 | 1.013 | 1.308 | 0.031 |

CHF: Congestive heart failure; HFpEF: Heart failure with preserved ejection fraction; HFrEF:

Kaplan-Meier curves of patients with no CHF, HFrEF, and HFpEF redemonstrated the significant difference in ischemic stroke rates between the three groups. We found that HFpEF patients had the highest stroke rate during follow-up, compared to both HFrEF patients and patients without CHF (Figure 1).

**Figure 1.**
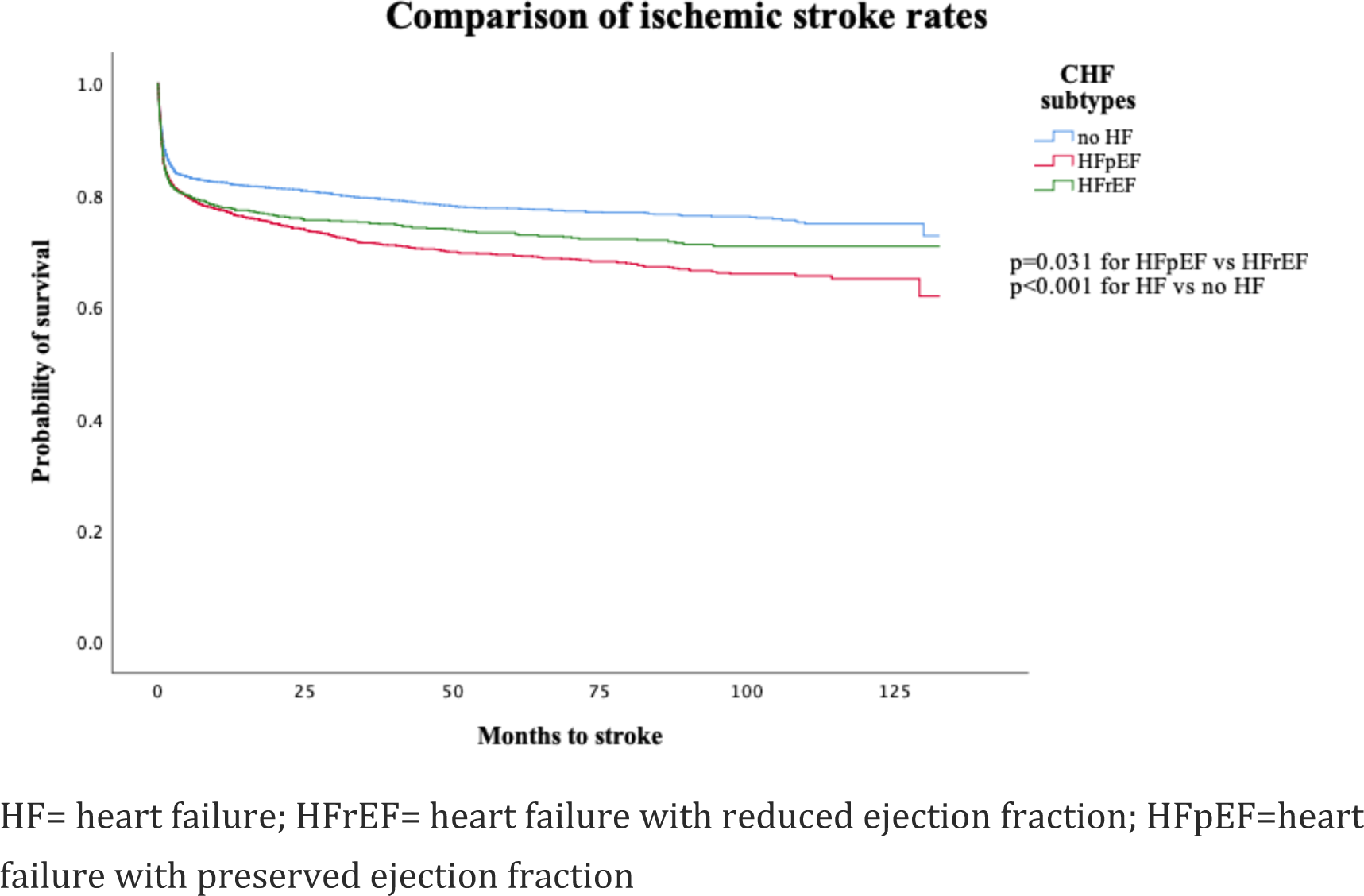
Comparison of ischemic stroke rates in patients with no heart failure, HFrEF, and HFpEF. During long term follow-up, HFpEF group had the highest rate of ischemic stroke, significantly higher than that of patients in HFrEF group (p=0.031).

### No difference in stroke risk between different subtypes of heart failure after adjustment for other risk factors

Because HFpEF patients had a higher CHA2DS2–VASc score or more comorbidities compared to HFrEF patients, it was possible that these risk factors might be confounding factors when evaluating ischemic stroke risk. We used Cox regression model to evaluate the stroke risks in different CHF subgroups to adjust for these risk factors.

After adjustment for other risk factors of stroke (components of CHA2DS2– VASc score including female sex, age, hypertension, diabetes mellitus, previous stroke, CAD, MI, and peripheral arterial disease), the risk of stroke between HFpEF and HFrEF was not significantly different (HR 1.001 (0.877-1.142), p=0.994, with HFrEF as reference). Among various known risk factors, history of stroke was associated with the highest increase in stroke incidence, with over 13-fold, followed by hypertension, with only 1.46-fold increasing risk (Table 3).

**Table 3.** Results of multivariate adjusted cox regression with other risk factors of stoke.

| Variables | HR | 95.0% CI for HR |  | p-value |
| --- | --- | --- | --- | --- |
|  |  | Lower | Upper |  |
| Sex (female vs male) | 1.102 | 1.007 | 1.206 | 0.035<sup>†</sup> |
| History of Stroke | 13.115 | 11.928 | 14.42 | <0.001<sup>†</sup> |
| Hypertension | 1.461 | 1.307 | 1.633 | <0.001<sup>†</sup> |
| Diabetes Mellitus | 1.107 | 1.01 | 1.213 | 0.03<sup>†</sup> |
| Coronary arterial disease | 1.074 | 0.975 | 1.183 | 0.15 |
| Myocardial infarction | 1.207 | 1.054 | 1.382 | 0.006<sup>†</sup> |
| Peripheral arterial disease | 1.153 | 0.989 | 1.345 | 0.07 |
| Age | 1.003 | 0.999 | 1.007 | 0.126 |
| No HF vs HFrEF | 0.836 | 0.737 | 0.948 | 0.005<sup>†</sup> |
| HFpEF vs HFrEF | 1.001 | 0.877 | 1.142 | 0.994 |

<sup>†</sup>: p-value<0.05; §: with HFrEF as reference

Dividing patients with CHF into three categories in regards to their CHA2DS2– VASc score, specifically with scores of 0-2, 3-5, and >5, we found that in each subgroup, there was no difference in ischemic stroke rate between CHF subtypes. Notably, over half of the patients were categorized in scores of 3-5, in which the Kaplan-Meier curves were closest (p=0.853) (Figure 2.)

**Figure 2.**
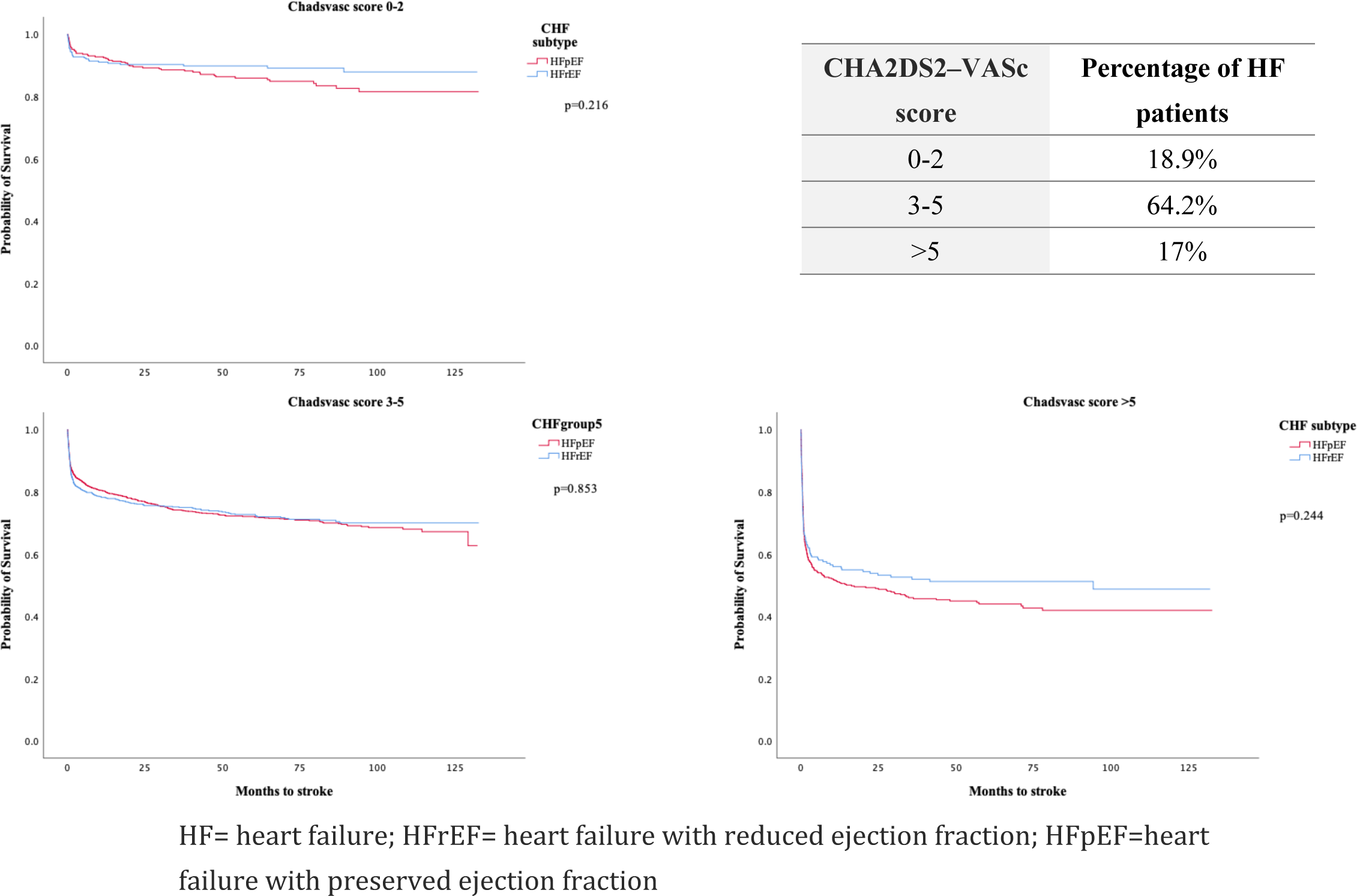
Comparison of ischemic stroke rates stratified by CHA2DS2–VASc score. When stratified by CHA2DS2–VASc score, the difference in ischemic stroke rate between HFrEF and HFpEF was no longer observed. (p=0.216, 0.853, 0.244 for CHA2DS2–VASc score 0-2, 3-5, and >5 respectively). Of note, the Kaplan-Meier curves were closest in the 3-5 category, in which had the majority of patients.

Our results show that both HFpEF and HFrEF were risk factors for stroke development in AF patients, and when taking into consideration other risk factors contributing to an ischemic stroke event, the two subtypes carry comparable increased risk.

## Discussion

This study represents the first long-term follow-up investigation comparing the roles of HFrEF and HFpEF in contributing to ischemic stroke risk in patients with AF in the Asia-Pacific region, and potentially worldwide.

In this real-world longitudinal follow-up study of AF patients with no CHF, HFpEF, or HFrEF, both HFpEF and HFrEF were associated with a higher risk of stroke. Specifically, HFpEF showed a greater risk when compared to HFrEF (HR 1.151 (1.013-1.308), p=0.031). Interestingly, we also observed a higher prevalence of previous stroke history in HFpEF patients compared to those with no CHF and HFrEF.

However, after conducting a multivariable analysis, the difference in stroke risk between HFpEF and HFrEF patients was no longer significant, and both groups exhibited around a 1.2-fold increased risk in stroke development. This was redemonstrated upon subgrouping patients in regards to their CHA2DS2–VASc score, with no difference in ischemic stroke risk between CHF subtypes in each subgroup.

The disparity in results before and after multivariable analysis could be attributed to the higher mean CHA2DS2–VASc score observed in HFpEF patients at the time of enrollment. This group of patients also had a greater prevalence of pro-inflammatory and metabolic comorbidities, such as diabetes mellitus and hypertension.

To the best of our knowledge, this study represents the longest follow-up investigation assessing the impact of different CHF subtypes in East Asian AF patients. Notably, our study cohort includes the largest number of CHF patients among Asian studies focusing on this subject.

### Development of CHA2DS2–VASc score

Current guidelines recommend the use of the CHA2DS2–VASc score as a risk stratification tool for predicting stroke risk and determining the need for anticoagulant therapy in patients with AF (3, 12, 13). Patients are categorized using a clinical evaluation approach that involves scoring based on the presence of several well-documented risk factors. The letter “C” in the CHA2DS2–VASc score represents congestive heart failure. As per the 2020 ESC guidelines, this includes clinical heart failure, encompassing those with recent decompensated heart failure regardless of ejection fraction, objective evidence of moderate to severe left ventricular dysfunction irrespective of symptoms, and hypertrophic cardiomyopathy (HCM) (3).

The criteria for the CHA2DS2-VASc score were initially defined and developed at a time when heart failure mainly referred to patients with reduced LVEF or left ventricular systolic dysfunction. However, it remains unclear and a continuous subject of debate whether these criteria are applicable to patients with HFpEF. In other words, it is uncertain whether the “C” in the CHA2DS2-VASc score should also be considered for this specific population. Further research is needed to address this question and provide clarity on the matter.

The CHA2DS2–VASc score is based on its predecessor, the CHADS2 score, further incorporating vascular disease, age of over 65 years old, and sex as modifying factors(14). Notably, the development of CHADS2 score was based on previous scoring models as well, derived from the Atrial Fibrillation Investigators (AFI) and the stroke prevention in atrial fibrillation (SPAF) trial. Information on the former was collected from five other trials, each with slightly different definitions of heart failure in their cohort, with none including data on ejection fraction(15). Current CHA2DS2– VASc score used the Euro Heart Survey on the AF population as their validation group, which defined heart failure as the “Onset of new heart failure, or worsening of known heart failure” (16, 17). Importantly, the term “heart failure” did not have a consistent definition, and data on LVEF was not included in these trials(10).

2022 AHA/ACC/HFSA Guideline for the Management of Heart Failure defines HFpEF as LVEF≥50%, with evidence of spontaneous or provokable increased LV filling pressures. Therefore, the presence of HFpEF without recent decompensation is not sufficient to meet the “C” criteria of the CHA2DS2–VASc score and require further clinical symptoms. This may be difficult to interpret as many HFpEF patients show hemodynamic abnormalities only during stress tests (18, 19). On the other hand, HFrEF itself is a guaranteed fulfillment of the condition. Our results show HFpEF and HFrEF have a similar increased risk in ischemic stroke after multivariable adjustments, thus both groups of patients may contain substantial thromboembolic risk.

The impact of ejection fraction on stroke risk has been an area of debate, with varying results. Banerjee et al. conducted a retrospective analysis of CHF patients with AF. Their findings revealed no significant difference in stroke incidence between AF patients with HFrEF and HFpEF, even after multivariable analysis. These results align with a recent meta-analysis on a similar topic, indicating that the stroke risk remains consistent across both CHF subtypes (20, 21). Another meta-analysis specifically investigating cardiovascular events in patients with AF found no significant difference in the occurrence of ischemic stroke between patients with HFpEF and HFrEF (22). These findings were replicated in a study conducted with Japanese patients hospitalized for CHF, which also produced similar results (23).

However, a study by McManus et al. demonstrated that pre-existing AF increased the risk of ischemic stroke specifically in patients with HFpEF (24). Recent data from a multicenter Korean registry supported these findings, revealing that the risks of ischemic stroke and thromboembolism were significantly higher in patients with HFpEF compared to those with HFrEF and heart failure with mid-range ejection fraction (HFmrEF) (11, 25).

This discordance in literature findings may be a result of several factors. Firstly, compared to previous studies with conflicting results, our CHF cohort consisted of a considerable number of patients, and the average follow-up duration was longer. As the effects of chronic underlying conditions may become significant in time, an extended follow-up time may be more beneficial in the observation of relevant risk factors. Compared to most studies, our study cohort had a greater proportion of HFpEF patients, which might have included patients in the lower severity spectrum. Recent studies show HFpEF patients compose the majority of heart failure patients and account for over 70% of elderly patients(26). The prevalence of AF is also seen to linearly increase with ejection fraction in patients hospitalized for acute CHF (27). As most registries include predominantly elderly patients, a lower number of HFpEF patients might have been biased with the exclusion of less severe patients, thus resulting in a worse prognosis. Furthermore, the cutoff value for HFpEF and HFrEF was 55% in our study, more stringent than most studies. In the 2016 ESC/Heart Failure Association (HFA) guidelines, LVEF≤40% is categorized as HFrEF, whereas LVEF≥50% is grouped as HFpEF(28). The grey area between HFpEF and HFrEF is referred to as HFmrEF. In the aforementioned studies, HFmrEF was found to pose less of a risk in stroke development compared HFpEF patients(11). Recent literature also showed similar stroke risk when using 40% and 50% as cutoff points for HFpEF and HFrEF patients (23). Furthermore, studies on heart failure patients showed HFmrEF patients to be responsive to therapies proven effective in HFrEF patients, suggesting the former may be a milder form of the latter (29, 30). Despite the above evidence, the possible inclusion of potential HFpEF patients in the HFrEF group only further strengthens our hypothesis, and is in line with our findings.

### Underlying comorbidities related to stroke

When compared to patients without CHF, both HFrEF and HFpEF had a higher prevalence of stroke-related risk factors and a higher incidence of stroke. This highlights the fact that regardless of the ejection fraction group, CHF itself is a risk factor for stroke. In fact, heart failure in non-arrhythmic patients is known to increase stroke incidence. Adelborg, Kasper et al. reported 1.5-to-2.1-fold increased risk of ischemic stroke in heart failure patients compared to those without during 30 years of follow-up in a population-based study (31). This was evident after adjustment for several risk factors including age, sex, and related comorbidities. Various studies show similar results, substantiating the relation (32, 33). Nevertheless, the direct cause and effect remain a topic of debate, as patients with CHF often carry multiple risk factors associated with stroke including hypertension, obesity, and diabetes mellitus.

While originally thought to be a spectrum of heart failure severity, HFpEF and HFrEF are now considered as two separate phenotypes, differing in risk factors, disease course, and management (34). Although sharing a portion of overlapping risk factors, HFrEF and HFpEF have also been shown to have differing predisposing conditions, with the former being a result of myocardial loss, and the latter commonly burdened with longstanding extra-cardiac comorbidities such as hypertension, type 2 DM, and older age (35). This difference is reflected in our cohort in which a prominent number of HFrEF patients with a history of MI and CAD, and a higher prevalence of hypertension in HFpEF patients.

Non-cardiac comorbidities are common in CHF patients, with past reports of up to 84% of HFrEF and 94% HFpEF patients with at least one (36). Notably, a higher prevalence of comorbidity burden has been shown in HFpEF patients of Asian ethnicity compared to their Western counterpart (37). Our study revealed a different patient profile in HFrEF and HFpEF groups, with the latter more predominately female, older, and hypertensive, as well as an overall higher mean CHA2DS2–VASc score. This was similar to previous reports in HFpEF patients with AF(11, 20, 22). Patient characteristics in our study are also comparable to previous reports in heart failure patients irrespective of arrhythmic state, which showed HFpEF patients to be predominantly female, older, and more often hypertensive(30, 38, 39). Indeed, evidence from cardiac structural changes in HFpEF patients have led to the hypothesis that hypertensive pressure overload and aging plays a major role in disease development in this groups of patients (40, 41, 42). Age, female sex and hypertension are notorious risk factors for stroke, and the increased prevalence of these conditions may indicate a greater benefit from anticoagulation therapy in this group.

In our cohort, the prevalence of MI and CAD were higher in the HFrEF group. This observation might be attributed to a shared etiology of post-MI and ischemic cardiomyopathy in this particular group. In contrast to HFpEF, in which development is attributable to chronic comorbidities, HFrEF has been shown to result from a distinct pathophysiological event, leading to a substantial and acute loss of myocytes, for example, myocardial infarction (35). Indeed, this is somewhat compatible with the previous results of Kamon, Daisuke et al., which showed 59% of AMI patients re-hospitalized for heart failure to have a LVEF of lower than 50%(43). Previous reports based on both AF and non-AF cohorts have consistently demonstrated a higher prevalence of MI and CAD in patients with HFrEF (11, 30, 36, 38). This may have contributed to the predominance of males in the HFrEF group as these patients may have had a greater risk of MI development (44). Overall, the complexity of heart failure patients plays a role in thromboembolic development, further contributing to mortality rates, and should also be taken into consideration in management.

### Limitations

This study was a longitudinal hospital-based investigation that recruited AF patients from a medical center, which might introduce inherent bias. Additionally, the treatment of heart failure in our patients could have influenced left ventricular function, further improving LVEF, thus potentially altering their CHF status during the follow-up period. Unfortunately, this aspect was not accounted for in the analysis. Moreover, although anticoagulant use was aligned with current guidelines, it was not stratified in our analysis and could have impacted the outcome of stroke.

Anticoagulant usage in our overall cohort was around 30%, which may have indicated a suboptimal prescription, influencing ischemic stroke rate. Nevertheless, as prevalence was similar across the three groups, it is unlikely that anticoagulation had a significant influence on our results.

### Conclusions

In conclusion, both HFrEF and HFpEF are risk factors for stroke in AF patients and should both be considered as “C” in the CHA2DS2–VASc score. With varying underlying patient characteristics, disparate management considerations for the two groups should be examined.

## Data Availability

Restrictions apply to the availability of some or all data generated or analyzed during this study to preserve patient confidentiality or because they were used under license. The corresponding author will on request detail the restrictions and any conditions under which access to some data may be provided.

## Acknowledgments

The authors thank all the participating individuals for their contribution to this study. The authors would like to express their thanks to the staff of Department of Medical Research for providing clinical data from National Taiwan University Hospital-integrated Medical Database (NTUH-iMD).

